# Words Matter: Political and gender analysis of speeches made by heads of government during the COVID-19 pandemic

**DOI:** 10.1101/2020.09.10.20187427

**Authors:** Sara Dada, Henry Ashworth, Marlene Joannie Bewa, Roopa Dhatt

**Author notes:** Corresponding author: Sara Dada.

## Abstract

**Background:** The COVID-19 pandemic has put a spotlight on political leadership and decision-making around the world. Differences in how leaders address the pandemic through public messages have practical implications for building trust and an effective response within a country.

**Methods:** We analyzed the public statements and speeches made by 20 heads of government around the world (Bangladesh, Belgium, Bolivia, Brazil, Dominican Republic, Finland, France, Germany, India, Indonesia, New Zealand, Niger, Norway, Russia, South Africa, Scotland, Sint Maarten, United Kingdom, United States, Taiwan) to highlight the differences between men and women leaders in discussing COVID-19 and pandemic response. We used a deductive analytical approach, coding speeches for specific themes based on language used and content discussed.

**Findings:** Five primary themes emerged across a total of 122 speeches on COVID-19, made by heads of government: economics and financial relief, social welfare and vulnerable populations, nationalism, responsibility and paternalism, and emotional appeals. While all leaders described the economic impact of the pandemic, women leaders spoke more frequently about the impact on local or individual scale. Women leaders were also more often found describing a wider range of social welfare services, including addressing to: mental health, substance abuse, and domestic violence. Both men and women from lower-resource settings described detailed financial relief and social welfare support that would impact the majority of their citizen population. While 17 of the 20 leaders used war metaphors to describe the COVID-19 virus and response, men largely used these with greater volume and frequency.

**Conclusion:** While this analysis does not attempt to answer whether men or women are more effective leaders in responding to the COVID-19 pandemic, it does provide insight into the rhetorical tools and types of language used by different leaders during a national and international crisis. This analysis provides additional knowledge on the importance and impact of political leaders speeches, messages, and priorities to inspire citizens adhesion to the social contract in the adoption of response and recovery measures.

## Background

The novel COVID-19 pandemic has shone a spotlight on political leadership and decision-making around the world. These decisions made by political leaders have critical implications for scientific research, vaccine development, healthcare delivery and systems, social and economic policy measures to contain the pandemic and ultimately for the health, well-being and life of citizens. In the current chaotic context, leadership and language matter; the ability of heads of governments and global health authority figures to communicate publicly on the impact of COVID-19 and the measures taken to mitigate risks are critical and closely scrutinized. Citizens’ perceptions, behaviors, and attitudes are significantly influenced by the type and quality of information or public services announcements to which they are exposed.^1^

Responses to the pandemic have varied significantly across countries and continents and are influenced not only by the magnitude of the pandemic, but also by pre-existing preparedness mechanisms and national leadership. For example, politicians’ statements are likely to have a powerful impact on citizen’s adherence to social distancing measures or mask-wearing. A recent study analyzing the context and chronology of presidential speeches in Brazil revealed the relationship between statements made by President Bolsonaro on COVID-19 prevention measures and adherence to social distancing policies.^2^ Additional research has noted that, while the difference is not statistically significant, countries led by women have seen better public health metrics in terms of COVID-19 response compared to countries led by men^3^. While risk perception and health behaviors may be non-partisan in theory, citizens’ behaviors are likely influenced by political leaders’ messages and calls to action or inaction.

At the time of writing, COVID-19 has not yet been eliminated in any but the smallest island states; some countries that had low early mortality rates are experiencing a resurgence and a second wave of infections is predicted before a vaccine is in widespread use. It would therefore be premature to conclude that some countries have done better in pandemic response than others since the picture may yet change. It is clear, however, that there are very different rhetorical styles across political leaders and that further work is needed to analyze how these influence public health messaging and public trust. In particular, it is worthwhile to consider if and how these differences are influenced by gender. This study analyzes the public speeches and statements made by heads of government in 20 countries around the world. The purpose is to understand the different language, rhetoric, and priorities expressed by men and women leaders in responding to the COVID-19 pandemic.

## Methods

### Setting

We set out to analyze official statements made by heads of government of the United Nations Security Council (Belgium, China, Dominican Republic, Estonia, France, Germany, Indonesia, Niger, Russian Federation, Saint Vincent and the Grenadines, South Africa, Tunisia, the United Kingdom, the United States, Vietnam), BRICS (Brazil, Russia, India, China, South Africa), and nation-states with women heads of government (Bangladesh, Barbados, Bolivia, Belgium, Denmark, Finland, Germany, Iceland, Namibia, New Zealand, Norway, Scotland, Serbia, Sint Maarten, Taiwan). We restricted the timeframe to February 26^th^ and April 6^th^ 2020 in order to capture the month of March, the beginning of when COVID-19 was spreading rapidly around the world and when international leaders began to respond actively. We recognize that gender is not binary, but for the purposes of this study have categorized political leaders as “man” or “woman” according to their assumed public gender identity, in the absence of other information on gender identity.

### Data collection

We searched for national addresses, statements, or speeches made to the public by all 29 countries’ heads of government (**Appendix 1**). Where fewer than two formal speeches were available, we also searched for press briefings (including only the introductory statements made by the head of government, before entertaining questions from the press), statements made to governing bodies (speeches to parliament), and other statements aimed towards the general public (podcasts, video announcements) and included these where available.

Written speeches were found on the public domain and via government websites. Where only a video was available, the speech was transcribed and cross-checked by a native speaker. All non-English speeches were translated into English using Google Translate and cross-checked by speakers fluent in the original language. In order to have a comparable final dataset, we aimed to include a range of geographic and political contexts that could be matched across the groups of men and women heads of government. We also aimed for gender parity and included the countries with the most available data in order to have a balanced pool. **Table 1**exhibits the final list of included countries and the number of statements made during the study period.

**Table 1:** Included countries and number of speeches available

| **Country** | **Leader** | **Man/**<br>**Woman** | **Region** | **Rationale for**<br>**inclusion** | **Number of**<br>**Speeches** |
| --- | --- | --- | --- | --- | --- |
| **Bangladesh<sup>4</sup>** | Hasina | Woman | Asia | Woman | 1 |
| **Belgium<sup>5</sup>** | Wilmès | Woman | Europe | UNSC, Woman | 3 |
| **Bolivia<sup>6</sup>** | Áñez | Woman | South America | Woman | 7 |
| **Brazil<sup>7</sup>** | Bolsonaro | Man | South America | BRICS | 4 |
| **Dominican Republic<sup>8,9</sup>** | Medina | Man | Caribbean | UNSC | 2 |
| **Finland<sup>10</sup>** | Marin | Woman | Europe | Woman | 3 |
| **France<sup>11</sup>** | Macron | Man | Europe | UNSC | 2 |
| **Germany<sup>12</sup>** | Merkel | Woman | Europe | UNSC, Woman | 4 |
| **India<sup>13</sup>** | Modi | Man | Asia | BRICS | 4 |
| **Indonesia<sup>14</sup>** | Widodo | Man | Asia | UNSC | 5 |
| **New Zealand<sup>15</sup>** | Ardern | Woman | Oceania | Woman | 3 |
| **Niger<sup>16</sup>** | Issoufou | Man | Africa | UNSC | 2 |
| **Norway<sup>17</sup>** | Solberg | Woman | Europe | Woman | 12 |
| **Russia<sup>18</sup>** | Putin | Man | Europe/Asia | BRICS, UNSC | 2 |
| **Scotland<sup>19</sup>** | Sturgeon | Woman | Europe | Woman | 14 |
| **South Africa<sup>20</sup>** | Ramaphosa | Man | Africa | BRICS, UNSC | 3 |
| **Sint Maarten<sup>21</sup>** | Jacobs | Woman | Caribbean | Woman | 12 |
| **Taiwan<sup>22</sup>** | Ing-wen | Woman | Asia | Woman | 2 |
| **United Kingdom<sup>23</sup>** | Johnson | Man | Europe | UNSC | 12 |
| **United States<sup>24</sup>** | Trump | Man | North America | UNSC | 25 |

### Analysis

Speech transcripts were uploaded and analyzed into NVivo. Two independent authors (SD, HA) used a grounded theory approach to analyze the speeches and coded themes that emerged in the data. A sample of six transcripts from different countries was used to develop the codebook applied to the rest of the dataset and was checked for data saturation.

### Ethical considerations

This research did not require an institutional board review approval. Data were collected online from publicly available and accessible speeches and did not involve any human subjects.

## Results

A total of 20 countries across six continents were included in this analysis, with 122 speeches analyzed. In the dataset of 122 speeches, 61 were delivered by women and 59 by men. **Table 2**provides some contextual information on the included countries, such as democracy index and gender development index scores and ranked by COVID-19 case count as of 30^th^ June 2020 (**Appendix 2**provides additional detail on COVID-19 context per country over time).

**Table 2:** Included countries ranked by COVID-19 case rate, including Democracy Index and Gender Development Index scores

| *Country* | *M/W* | *Cases Per Capita as of 30 June<sup>25</sup>* | *Democracy Index 2019<sup>26</sup>* | *Democracy Index* | *Gender Development Index 2018<sup>27</sup>* | *Gender Development Index Group* |
| --- | --- | --- | --- | --- | --- | --- |
| *United States* | M | 0.7667% | 7.96 | Flawed | 0.991 | 1 |
| *Brazil* | M | 0.6324% | 6.86 | Flawed | 0.995 | 1 |
| *Belgium* | W | 0.5300% | 7.64 | Flawed | 0.972 | 2 |
| *United Kingdom* | M | 0.4595% | 8.52 | Full | 0.967 | 2 |
| *Russia* | M | 0.4439% | 3.11 | Authoritarian | 1.015 | 1 |
| *Scotland* | W | 0.3473% | N/A | N/A | N/A | N/A |
| *Dominican Republic* | M | 0.2933% | 6.54 | Flawed | 1.003 | 1 |
| *Bolivia* | W | 0.2701% | 4.84 | Hybrid | 0.936 | 3 |
| *South Africa* | M | 0.2432% | 7.24 | Flawed | 0.984 | 1 |
| *France* | M | 0.2404% | 8.12 | Full | 0.984 | 1 |
| *Germany* | W | 0.2319% | 8.68 | Full | 0.968 | 2 |
| *Sint Maarten* | W | 0.1796% | N/A | N/A | N/A | N/A |
| *Norway* | W | 0.1633% | 9.87 | Full* | 0.990 | 1 |
| *Finland* | W | 0.1301% | 9.25 | Full | 0.990 | 1 |
| *Bangladesh* | W | 0.0861% | 5.88 | Hybrid | 0.893 | 5 |
| *India* | M | 0.0411% | 6.90 | Flawed | 0.829 | 5 |
| *New Zealand* | W | 0.0244% | 9.26 | Full | 0.963 | 2 |
| *Indonesia* | M | 0.0201% | 6.48 | Flawed | 0.937 | 3 |
| *Niger* | M | 0.0044% | 3.29 | Authoritarian | 0.298 | 5 |
| *Taiwan* | W | 0.0019% | 7.73 | Flawed | N/A | N/A |

Five major themes emerged across speeches: the economy and financial relief, social welfare and vulnerable populations, nationalism, responsibility and paternalism, and emotional appeals. While speeches and statements covered a wide range of content, these themes were where differences in priorities and language across countries and across leaders were most distinct. Our final interrater reliability, Kappa score, was 0.81, suggesting moderate to strong agreement amongst coders^28^.

### The Economy & Financial Relief

The economy was a central theme across leaders regardless of geography, level of gross domestic product, or gender. Eighteen of the 20 leaders, excluding India’s Modi and Bangladesh’s Hasina, specifically detailed the programs that would provide financial relief. Seven leaders (3 women - Bolivia, Germany, Norway; 4 men - France, Indonesia, Russia, USA) focused on small businesses across 20 speeches, while six leaders (3 women - Germany, Norway, Scotland; 3 men - Dominican Republic, South Africa, USA) discussed large businesses across 21 speeches. Informal economies were discussed by three leaders (1 woman - Norway; 2 men - Dominican Republic, South Africa) in five speeches.

All leaders were acutely aware of and highlighted the devastating economic impacts that COVID-19 would have on their nations. Almost all leaders made reference to financial support for families, small businesses, and larger corporations. However, there was a difference in how men and women addressed this economic impact. Despite the near equal number of speeches with economic references made across genders, this analysis demonstrates that women leaders tended to prioritize discussing the economy at the level of individuals and small businesses, while their men counterparts more often focused on larger businesses and corporations (**Table3**). Besides emphasizing economic support for individuals and employees, a woman (Scotland’s Sturgeon) was the only leader to discuss labor unions. This connects to a larger theme across categories that women demonstrated a more people-centered approach compared to the men.

**Table 3:** Leaders’ references to small versus big businesses

| *Country* | *Number of References to Small Businesses* | *Number of Speeches (of total speeches)* | *Average Reference Coverage per speech* | *Number of References to Big Businesses* | *Number of Speeches (of total speeches)* | *Average Reference Coverage per speech* |
| --- | --- | --- | --- | --- | --- | --- |
| *Bolivia* | 5 | 3 (of 3) | 5.30% | -- | -- | -- |
| *Dominican Republic* | -- | -- | -- | 2 | 1 (of 2) | 1.77% |
| *France* | 1 | 1 (of 2) | 0.27% | -- | -- | -- |
| *Germany* | 1 | 1 (of 4) | 10.39% | 2 | 1 (of 4) | 5.28% |
| *Indonesia* | 1 | 1 (of 4) | 3.03% | -- | -- | -- |
| *Norway* | 4 | 2 (of 12) | 4.05% | 8 | 3 (of 12) | 5.03% |
| *Russia* | 2 | 1 (of 2) | 2.32% | -- | -- | -- |
| *Scotland* | -- | -- | -- | 3 | 2 (of 14) | 3.34% |
| *South Africa* | -- | -- | -- | 2 | 1 (of 3) | 1.13% |
| *United States* | 28 | 14 (of 25) | 2.69% | 55 | 12 (of 25) | 8.05% |

Of the men who discussed economic support to smaller businesses, American President Donald Trump and French President Emmanuel Macron stood out. Macron specifically focused on providing unconditional support to business owners. On the 12th of March he stated:

> *“We will not add the fear of bankruptcy for entrepreneurs, concerns about unemployment and the challenge of making ends meet at the end of the month to health worries. Every effort will therefore be made to protect our employees and to protect our companies, regardless of the cost”.^11^*

Trump frequently discussed supporting small businesses, and referenced them in 14 of his 25 speeches. However, these references were few compared to the emphasis he put on addressing large corporations and CEOs. The first topic Trump discussed in half of his first ten speeches was the economic aspects of COVID-19, rather than the medical or health repercussions for society. Men leaders tended to focus more on supporting large business and emphasizing overall national economic recovery over individual support. This either came in subtle forms like Brazilian President Jair Bolsonaro stating on the 31st of March, *“We have a mission: to save lives, without leaving jobs behind”^7^*or in more overt forms such as Trump directly naming and thanking companies and inviting their CEOs to speak at formal press briefings, particularly on the 13^th^, 15^th^, and 30^th^ of March. Trump continually emphasized his cooperation and support of larger businesses, and flagged how economic support was coming to ensure that they would not suffer.

Women leaders who discussed supporting small businesses highlighted the importance of protecting workers as well as individuals and families. Bolivia’s interim President Jeanine Áñez, described on the 15th of March how programs for *“small and medium-sized entrepreneurs who are suffering and who will suffer due to this health crisis”* would also *“help Bolivian families who will suffer an economic impact from the coronavirus.”^6^* In Scotland, First Minister Nicola Sturgeon stated in an address on March 22nd, *“To the vast majority of employers I say this – the solution to the challenges that I know you are facing now is not key worker status. It is new shift patterns, it is working from home, it is dropping non-essential tasks. And that is what you can do to help all of us save lives.”^19^*Women leaders were also specific about their economic support for small businesses. Finland’s Prime Minister Sanna Marin and Germany’s Chancellor Angela Merkel both detailed financial programs and stimulus funds for small businesses.

### Vulnerable Populations & Social Welfare

German Chancellor Merkel described the relationship between the economic and social impacts of the pandemic best in a speech on the 23rd of March with the statement, *“many are experiencing a social emergency due to the economic impact*.”*^12^*The COVID-19 pandemic has catalyzed new social emergencies and deepened chronic challenges including social inequities. While the two are intertwined, distinct vulnerable populations may require distinct forms of social welfare. Just as the response to economic concerns varied across leaders, so did their response to supporting new and longstanding vulnerable populations through different forms of social welfare in their speeches.

With regards to specific vulnerable populations, only women heads of government noted the vulnerability of migrants and refugees, individuals with mental health and substance use issues, and victims of domestic violence. When women leaders spoke about and to these populations, they did so with strong, empathetic and persuasive language. For example Bolivia’s Áñez specifically called out and condemned all acts of violence perpetrated against women:

> *We have received many complaints about sexist violence and domestic violence during these days of quarantine. I want to be very clear at this point. We are going to fall with the full weight of the law before those who commit violence against women and against families. And they already know me. I am going to be firm, I am going to be firm in the defense of Bolivian women and families^6^*

Immigrants, asylum seekers, and refugees have heightened vulnerability in this pandemic. The only leaders to mention these populations, and describe support for them were Belgium’s Prime Minister Sophie Wilmès and New Zealand’s Prime Minister Jacinda Ardern. For example on the 5th of April, Wilmès stated, “*many decisions have also been taken in the field of mobility, asylum and support for the most vulnerable people. These decisions were taken by the federal government as a team, in collaboration with parliaments and the social partners.”^5^*

Men and women both mentioned other vulnerable populations including children, the sick or immunocompromised, and the elderly consistently across regions. These references were both in relation to how collectively each country needed to take care of these populations, and also the specific actions each government was taking. Typical comments include ones from Johnson on the 12th of March, “ *The most important task will be to protect our elderly and most vulnerable people during the peak weeks when there is the maximum risk of exposure to the disease and when the NHS will be under the most pressure’’^23^*and Sturgeon on the 17^th^ of March, *“Firstly, people who are over 70, second people with underlying health conditions for which they get the flu vaccine, and third women who are pregnant. We are strongly advising them to stay at home as much as possible, and to significantly reduce unnecessary social contact.”^19^*Some leaders particularly stood out including Norwegian Prime Minister Erna Solberg’s focus on children, which included a press briefing specifically for children.

In terms of social welfare, all men and women leaders mentioned unemployment, healthcare costs and access, food, and housing. Leaders of low and middle-income countries stressed access to food. Examples include Bangladeshi Prime Minister Sheikh Hasina on the 25th of March, *“Assistance will be provided to low-income people in their respective villages under the ‘Homecoming’ program. Free house, 6 months food and cash will be provided to the homeless and landless”^4^*and South African President Cyril Ramaphosa on the 30th of March, “ *The elderly and the frail need people to care for them. Some of those who live on the streets are without shelter or food.”^20^*

Women leaders dove to a deeper depth on social welfare particularly related to the vulnerable and need for social support. This was seen in how their mentioning less traditional, yet equally necessary forms of social welfare including day care, mental health, and support for victims of domestic violence and sexual assault. Only Áñez and Sturgeon directly addressed domestic violence and measures to combat the reports of increased incidence during the pandemic. Another example is Sturgeon’s comprehensive plan to provide equitable support for mental health described on the 27th of March:

> *We are putting an additional £3.8 million immediately into the NHS’s mental health support services. £0.5 million of that will support for the Breathing Space phone line and web service, and £2.1 million will be for the NHS’s Mental Health Hub… The mental health hub will expand its staffing in the coming weeks, so that it can become available to the public 24 hours a day, 7 days a week.^19^*

This focus was consistent across most women leaders and can be seen in how Sint Maarten’s Prime Minister Silveria Jacobs mentioned it in over half of her speeches, often with specific actions such as on the 20th of March, “*designate the first hour of business to senior citizens, as well as vulnerable persons such as pregnant women and persons with children. This is in order to minimize contact.”^21^*

### Nationalism

Reacting to a pandemic in a globalized society brings up an interesting dichotomy of national versus international interests. In our analysis the theme of nationalism appeared in a number of ways in leaders’ responses. These include actions such as enforcing border control and rhetoric such as blame, advocacy for global cooperation, and urging a sense of patriotic duty and sacrifice.

While only three leaders incorporated a tactic of blame in their speeches (Áñez, Bolsonaro, and Trump), each did so to a different extent. Trump blamed a third party in 14 of his 25 speeches. However, in only one speech on the 15^th^ of March, Áñez said *“we receive a very neglected and flawed health system. So now our effort must be double.” ^6^*This reflects a similar theme in Trump’s speeches, such as on the 5^th^ of April when he stated: *“And you remember, we inherited a broken system.”^24^*However, the biggest target of Trump’s blame remains China. He is the only leader to consistently refer to COVID-19 as *“the Chinese virus,” ^24^*rather than using its official or scientific name. On the 14^th^ of March, he stated: *“It’s something that nobody expected. It came out of China, and it’s one of those things that happened.” ^24^*Beyond this, Trump and Bolsonaro both blame China and the media for their roles in the pandemic. On the 24^th^ of March, Bolsonaro accused the media of stirring up panic in the country: *“Considerable part of the media went against the grain. They spread exactly the feeling of fear, with the announcement of the large number of victims in Italy as their flagship.”^7^*

While leaders of 13 countries (8 women – Bangladesh, Bolivia, Finland, Germany, New Zealand, Norway, Sint Maarten, Taiwan; 5 men – Dominican Republic, France, Niger, South Africa, USA) discussed limited international borders or suspending flights to or from specific regions, Trump stressed travel restrictions taking an isolationist approach. On the 24^th^ of March he emphasized the need for self-reliance with:

> *We should never be reliant on a foreign country for the means of our own survival. I think we ‘ve learned a lot. We ‘ve learned a lot. This crisis has underscored just how critical it is to have strong borders and a robust manufacturing sector. . . America will never be a supplicant nation. We will be a proud, prosperous, independent, and self-reliant nation. We will embrace commerce with all, but we will be dependent on none.^24^*

This mentality contrasted the emphasis several other leaders placed on global cooperation. Four nations led by men discussed the value and importance of global cooperation (France, Niger, UK, USA), while seven women-led nations did the same (Bangladesh, Bolivia, Finland, Germany, Norway, Sint Maarten, Taiwan). Even amongst the men who did discuss global cooperation, they discussed it far less, with just one mention each, except for Trump who on three occasions described working with other nations referring to how America was *“helping other nations - many other nations; we ‘re helping them a lot.”^24^*On the 12^th^ of March, Macron stressed “ *This virus doesn’t have a passport. We need to join forces, coordinate our responses and cooperate*”^11^ and on the 17^th^ Johnson simply said *“while we need national unity, we also need international cooperation.” ^23^*On the 27^th^ of March, Niger’s President Mahamadou Issoufou highlighted the role of globalization:

> *I appeal to international solidarity to support the implementation of this plan. This solidarity is more than ever justified because the increase in the speed of means of transport has canceled the distances between regions of the world and between countries*.. . *More solidarity and more equality, this must be the rallying cry, this must be the new creed. We must share both the risks and the benefits of globalization.^16^*

When considering the women, Marin, Merkel, and Solberg stressed collaboration across the European region and Áñez emphasized cooperation with the World Health Organization. Sint Maarten, as an overseas territory of the Netherlands and an island joined to a French territory of similar name, has a unique political context that explains Jacobs’s common references to Dutch and French and European aid in response. Taiwan has a similarly unique political context that could explain President Ing-wen’s calls for *“international responsibility [and] full international cooperation.”^22^*

A rhetorical tool used across leaders was the emphasis on patriotic duty to encourage sacrifice. Twelve countries (7 men – Brazil, Dominican Republic, France, India, South Africa, UK, USA; 5 women – Bolivia, New Zealand, Norway, Scotland, Taiwan) used language calling on the public to love their country, serve their nation, or come together as one. In 11 countries, this went a step forward and called for self-sacrifice in the name of patriotism or service to the greater common good (5 men – Dominican Republic, France, South Africa, UK, USA; 6 women – Belgium, Bolivia, New Zealand, Norway, Scotland, Taiwan). Notably, some of the women emphasized that this sacrifice is made for others – such as Wilmès’s statement on the 5^th^ of April: *“The sacrifices we make are very great, especially for people who are alone” ^5^*or Solberg on the 12^th^ of March: *“We must put life and health first anyway. For each other. And for all those we love.”^17^*Trump stressed sacrifice for the nation as a whole on at least six occasions, including the 18^th^ of March: *“Every generation of Americans has been called to make shared sacrifices for the good of the nation.” ^24^*Johnson, Macron, Medina, and Ramaphosa focused on acknowledging sacrifices or expressing gratitude for the sacrifices made by all citizens and specific subgroups, such as healthcare providers.

### Responsibility vs. Paternalism

Leaders included in this analysis often conveyed the importance of citizens taking actions to protect themselves and adhere to governmental guidelines. This analysis highlights two forms of emotional appeal used to convey this message: responsibility and paternalism. We defined responsibility as rhetoric that encouraged individuals to act independently to adhere to guidelines and paternalism as rhetoric that employed tactics such as shame, guilt, or punishment to influence the desired behavior. Both men and women leaders used these tactics equally, without a significant difference by gender. We found responsibility used by 15 of the leaders (8 men – Dominican Republic, France, India, Indonesia, Russia, South Africa, United Kingdom, United States; 7 women – Belgium, Bolivia, Germany, New Zealand, Norway, Scotland, Sint Maarten) and paternalism used by 12 (6 men – Dominican Republic, France, India, Indonesia, South Africa, United States; 6 women – Belgium, Bolivia, Germany, New Zealand, Scotland, Sint Maarten)

Leaders often balanced the use of paternalism and responsibility within a speech. One example of this was seen in Ardern’s speech on March 23rd. Initially she used more paternalistic language to emphasize the importance of adhering to guidelines:

> *That’s why sticking to the rules matters. If we don’t – if you hang out with that friend at a park or see that family member for lunch, you risk spreading COVID -19 and extending everyone’s time in Level 4. . . Failure to play your part in the coming days will put the lives of others at risk. There will be no tolerance for that and we will not hesitate in using enforcement powers if needed.^15^*

This quote uses guilt of spreading COVID-19 and extending government restrictions as a means of motivating compliance. She later concludes her speech with responsibility, particularly focusing on empowering citizens to recognize the important role they play:

> *You may not be at work, but that doesn’t mean you don’t have a job. Your job is to save lives, and you can do that by staying home, and breaking the chain. . .And finally, if you have any questions about what you can or can’t do, apply a simple principle. Act like you have COVID-19. Every move you then make is a risk to someone else. That is how we must all collectively think. That’s why the joy of physically visiting other family children, grandchildren, friends, neighbours is on hold. Because we ‘re all now putting each other first. And that is what we as a nation do so well. So New Zealand, be calm, be kind, stay at home. We can break the chain.^15^*

Other examples of responsibility include addressing the importance of personal responsibility and taking ownership of the pandemic. This was displayed by South Africa’s President Cyril Ramaphosa on March 30th, *“Let us not make the mistake of thinking this is somebody else’s problem;”^20^*Belgium’s Prime Minister Sophie Wilmès on March 19th, *“Each of us has a role to play. Not only to protect ourselves, but also to protect our loved ones and our fellow citizens*”;^5^ and United Kingdom’s Prime Minister Boris Johnson on March 20th, *“Each and every one of us is now obliged to join together. To halt the spread of this disease. To protect our NHS and to save many, many thousands of lives.”^23^*Here responsibility highlights social contracts and the duty to protect other citizens. Paternalism was often partnered with language around the enforcement of policies by other citizens or the government, as used by Ardern. Examples include Indonesian President Joko Widodo on March 20th, *“Don’t hesitate to reprimand those who are not disciplined in keeping the distance, washing their hands, and maintaining their health”^14^*and Sturgeon on March 22nd *“My message to them is close now. We will have emergency powers within days to force you to close and we will use these powers if we have to. But you should not wait for that.”^19^*Collectively, paternalism and responsibility were important tools of emotional rhetoric used by all leaders to motivate their citizens to adhere to newly implemented guidelines.

### Emotional Appeals

A common rhetorical tool throughout the pandemic, and even across infectious disease discourse, is the use of war analogies – likening an outbreak to a war. Ten of the included leaders specifically describe the pandemic as a “war” or “battle (5 women - Bangladesh, Bolivia, Norway, Scotland, Taiwan; 5 men - Dominican Republic, France, India, United Kingdom, United States). This number expands to 17 when we consider the use of *any*war rhetoric such as words along the lines of “fighting” and “enemy.” As seen in **Table 3**, while this metaphor is used across genders, the frequency and aggression of these analogies seems to be stronger with the men. In the 19 speeches made by women who used war metaphors, they average 6.1 references to this rhetoric. In the 40 speeches made by men with this language, they average 25.4 references to war metaphors. For example, Trump used war analogies 136 times across 23 speeches, Johnson 15 times across 7 speeches, Modi 30 times across all 4 of his speeches. Meanwhile, the only woman to use this rhetoric in double digits was Áñez with 26 occurrences in her 5 speeches. On average, for speeches that did employ war rhetoric or aggressive language, it constituted 3.5% of each speech made by men, and 2.8% for the women (1.9% without Áñez).

**Table 4:** Leaders’ use of war metaphors across speeches

| *Country* | *Number of References* | *Number of Speeches (of total speeches)* | *Average Reference Coverage per speech* |
| --- | --- | --- | --- |
| *Bangladesh* | 3 | 1 (of 1) | 3.04% |
| *Belgium* | 9 | 3 (of 3) | 3.19% |
| *Bolivia* | 26 | 5 (of 7) | 10.00% |
| *Dominican Republic* | 1 | 1 (of 2) | 0.61% |
| *Finland* | 1 | 1 (of 3) | 1.37% |
| *France* | 9 | 1 (of 2) | 3.65% |
| *India* | 30 | 4 (of 4) | 5.39% |
| *Indonesia* | 2 | 1 (of 4) | 1.96% |
| *New Zealand* | 4 | 2 (of 3) | 2.21% |
| *Niger* | 7 | 2 (of 2) | 3.95% |
| *Norway* | 4 | 2 (of 12) | 1.60% |
| *Scotland* | 4 | 2 (of 14) | 1.29% |
| *Sint Maarten* | 1 | 1 (of 12) | 0.59% |
| *South Africa* | 2 | 1 (of 3) | 1.49% |
| *Taiwan* | 3 | 2 (of 2) | 1.87% |
| *United Kingdom* | 15 | 7 (of 12) | 4.76% |
| *United States* | 136 | 23 (of 25) | 4.51% |

The language around war ranged in describing the battles, enemies, and weapons employed in the fight against COVID-19. On 16^th^ March, Macron states: *“We are at war, admittedly a health war: we ‘re fighting neither an army nor another nation. But the enemy is there, invisible, elusive, and it’s making headway.”^11^*Four other leaders also describe the virus as an *“invisible enemy”^16,17,23,24^*(Issoufou, Johnson, Solberg, and Trump) and Johnson even declares it *“the invisible killer”^23^*on the 23^rd^ of March, just days before announcing his own illness. Issoufou and Ardern equate options for response to the pandemic to weapons. For example, on the 17^th^ of March Issoufou explains *“The only weapon that exists today is prevention.”^16^*

Bolivia’s Áñez was the woman leader with the most war references, for example expressing on the 15^th^ of March: *“I will dedicate 100% of my time to fighting for the health of Bolivians. It will be a tough battle and it will be a long one but I want you to know that it is a battle that we are going to win if we do it among all Bolivians.” ^6^*In observing Áñez’s use of this language, it is relevant to consider the context of her political power as well. In addition to being the only woman in this dataset to reference war in the double digits, she is also the only leader in this pool who came to power after a coup. It is worth noting how the political and societal context of the country, Bolivia, could influence the language used by their head of government. Notably, both times Ing-wen employs the war metaphor, it is to describe the necessity for *“full international cooperation [as] the only way to ensure that the international community can win this battle.”^22^*In the case of Ing-wen, it is worth noting that Taiwan is the smallest state included in this analysis and it is possible that its contested relationship with China influences views on globalization and international cooperation.

Trump is not the only leader to utilize this blasé tone. This is another similarity amongst a group, though not all, of the men leaders. In Brazil, India, the UK, and the US, heads of government often dismissed the severity and concern over COVID-19. On the 24^th^ March, Bolsonaro explained, *“In my particular case, due to my athlete’s history, if I was infected by the virus, I would not have to worry, I would not feel anything or I would be, at most, suffering from a light Hu or a light cold, as the well-known doctor from that well-known television said*.*”^7^* This personal nonchalance was clear in another speech a week later when he said *“The coronavirus came and one day it will go away, unfortunately we will have losses along the way.”* Similarly, Modi and Johnson stress that *“everything is okay”^13^*and the need for *“going about our business as usual,” ^23^*respectively. This same attitude is demonstrated in the way Trump discusses the voluntary nature of wearing face masks on the 3^rd^ of April: *“So with the masks, it’s going to be, really, a voluntary thing. You can do it. You don’t have to do it. I’m choosing not to do it, but some people may want to do it, and that’s okay. It may be good. Probably will. They’re making a recommendation. It’s only a recommendation. It’s voluntary.” ^24^*

This approach is a stark contrast to the language employed by some of the women leaders in this analysis. It was more common for women to stress empathy and compassion in their speeches to the public. One poignant example is Norway’s press briefings held specifically for children. On multiple occasions, Solberg held these events to answer the questions and concerns of the nation’s children, including on the 16^th^ of March when she said: *“Many children find this scary. I understand that well. It’s allowed to get a little scared when so many big things happen at once. It is allowed to be a little scared to get infected by the coronavirus.”^17^*Women leaders such as Áñez and Ardern also empathized with parents and families by stressing their experiences as mothers. While the men leaders included in this analysis also have children, none of them mentioned their roles as fathers or experiences with family. In the case of Scotland’s Sturgeon, this empathy included messages to the families who have lost loved ones to COVID-19 during the pandemic. Sturgeon began every speech with acknowledging the new deaths or hospitalizations in the country and explicitly sharing her condolences with those families and the public as a whole. She does this with honesty and compassion, for example on the 5^th^ of April with, *“The figure I will report tomorrow, is likely to be artificially low – though of course, each one matters and is a source of sadness to family and friends but also to me.”^19^*

Women leaders including Áñez, Ardern, Jacobs, Merkel, and Sturgeon called on their constituents to employ this empathy in their adherence to various restrictions and policies that may have been implemented. On the 19^th^ of March, Merkel stressed: *“These are not just abstract numbers in statistics, but this is about a father or grandfather, a mother or grandmother, a partner – this is about people. And we are a community in which each life and each person counts.”^12^*Áñez used similar language on the 25^th^ of March, asking the audience to think of their parents and grandparents who took care of them in the past and needed to be protected now. Ardern and Sturgeon emphasize a need to *“be kind”^15,19^*and Jacobs implores society to *“show love and caring to one another.”^15^*

The strongest emotional appeal from the men leaders came from the Dominican Republic’s now Former President Danilo Medina. Both of his speeches in March included vivid calls for solidarity and compassion, and on the 17^th^ of March he concluded his speech with:

> *It is true, we are facing the most serious public health alert in recent decades, but we are also facing one of the moments in history that shows us the true greatness of human beings. Being a Dominican has always meant facing challenges with courage and facing the future with optimism and this time will be no exception. Let’s not let fear cloud our gaze. Let us divest ourselves of all selfishness and see in each compatriot a member of our great family. Let us act with temperance, with rationality and always thinking about what is really important, what should unite us now, what we all want to preserve: the health of our grandparents and parents, of all those who are now more vulnerable and, of course, of our sons and daughters.^9^*

## Discussion

The purpose of this study is to analyze how a leaders’ gender may relate to their communication style during the initial response to the COVID-19 pandemic, rather than to declare whether countries led by men or women managed the early months of the pandemic more effectively. In this study, we specifically focus on the content of communications by heads of government, their communication style, and the differences in these approaches. It is important to note that these differences are influenced by more than just gender – such as geographic context, political ideologies, socio-economic context, political history of the country, and population demographics. However, our analysis shows that there are notable differences between men and women heads of government with practical implications for pandemic response.

This analysis highlights noticeable differences between the content men and women leaders used related to economics and social welfare and how they made emotional appeals to the public. While all leaders acknowledged the economic impacts of the pandemic, women were more likely to cite the impact on the micro-scale, emphasizing the impact on individuals and families. This continued in the way men and women leaders described social welfare mechanisms versus financial relief. In particular, only women acknowledged the impact of the pandemic on unique vulnerable groups including migrants and refugees, substance abusers, people with mental health challenges, and its impact on family care responsibilities and domestic violence. While we did not assess governments’ actual programmatic responses or policies, the focus on vulnerable populations by women heads of government demonstrates the prioritization of vulnerable populations and social cohesion in response.

Previous studies have described how the message-framing of leaders can influence decision-making processes and behavior on micro and macro scales – from vaccination behavior to trust in outbreak preparedness.^2,29-31^ According to the agenda-setting research paradigm^32^, messages highlighted in media or by leaders influence the selection and prioritization of issues in society as well as policy-agenda-setting. In fact, messages shared by opinion leaders through media are more likely to impact policy both by shaping public perception of risk and by shaping policy makers’ perception of public opinion. Government communication style is therefore likely to be associated with perceptions, responses to health threats, resulting in health, social, and economic outcomes for the public and nation.^33^ For example, President Obama’s use of the term “epidemic” rather than “outbreak” during the 2014-2016 West Africa Ebola epidemic transformed the language being used by the media to describe the growing emergency.^34^ This transition of terminology went beyond semantics, affecting the conceptualization of the crisis and therefore the resulting response.

Another clear difference across genders was the varying approaches in making emotional appeals. Across the board, men tended to use more war rhetoric while women tended to employ personal or empathetic appeals. Language that uses imagery of battles and threatening enemies can be a powerful tool to invoke fear in the audience.^35^ As examples, Prime Minister Johnson explicitly stated that *“we must act like any wartime government and do whatever it takes to support our economy”^23^*and President Trump demonstrated this very clearly in discussing the virus and the government’s response. Times of war require unprecedented action from the government in order to protect the populace. However, the fear and sense of urgency invoked by this language can also be used as a justification for a lack of transparency.^36,37^

Approaching the COVID-19 pandemic as a war has a strong influence on response mechanisms and proposed policies. In times of crisis, those in charge may be quick to fall back on existing structures of response that are not transparent.^38^ The decisions made in these times, particularly in attempting to mitigate a disease outbreak, will invariably affect and rely on the public. Yet, when decisions are taken behind closed doors, how they communicated to the public impacts on public trust and compliance with public health measures. It is precisely and especially during emergencies that the public requires transparent procedures and clearly communicated decisions. Building trust and enabling accountability depend on it, underpinning not only an efficient outbreak response but also an effective health system.^39^Accountability and transparency are critical to earn the public’s trust and shared information must be evidence-based. Elected officials must have some level of understanding of health and risk communication in order to communicate effectively in times of health crises, as well as the skills to translate science into policy to ensure evidence-based priorities and policy implementation.^40^

The empathetic and personal appeals that women made focused on compassion and social cohesion, such as Chancellor Merkel’s comment, “*these are not just abstract numbers in statistics, but this is about a father or grandfather, a mother or grandmother, a partner – this is about people. And we are a community in which each life and each person counts*.” Similar comments asking for compassion for others were seen consistently throughout speeches made by women. Commentators have picked up on these emotional appeals as being vital for generating social cohesion to generate a unified public response.^41^ In particular, Prime Minister Ardern has been hailed for her ability to generate trust through transparency and action through the social cohesion she inspires.^42^ Empathic statements and war rhetoric may both inspire a form of unity, but they have important practical differences. While war rhetoric plays to a collectivism based upon fear and division, empathy appeals to a collectivism based upon compassionate social cohesion.^43^ A fear-based approach may instigate conflict and marginalization.^44,45^

### Limitations

There are several limitations to this study. As with qualitative research, there is a limit to the comparability and generalizability of the data analyzed. This study relied on publicly available data that could be transcribed or translated to English with limited resources, which also made some speeches subject to errors in transcription and translation. Due to this and other differences amongst countries, the number of speeches from each country varies greatly. However, it is worth noting that there were roughly the same amount of speeches from men (n=59) and women (n=61) included. In identifying gender we did not directly ask heads of government for their gender identity, but instead relied upon gender identities reported in the news and through pronouns in speeches. We acknowledge, however, that gender is complex, self-identified, non-binary, and socially produced. Finally, countries have been impacted differently by the pandemic (incidence, prevalence, mortality rate) and have had different experiences with managing public health emergencies, which can affect their response, priorities, political and risk communication styles. This study did not consider this historical context nor the societal and political ideals and values that influence individual leaders, the language they use, and political priorities.

## Conclusion

While this analysis does not attempt to answer whether men or women are more effective leaders in responding to the COVID-19 pandemic, it does provide insight into the rhetorical tools and types of language used by different leaders during a national and international health crisis. This language matters because it influences how leaders inspire citizens’ compliance with response and recovery measures. This analysis explores how men and women heads of government tend to use different types of language. There are certainly exceptions, but this study points to how imperative communication is in shaping a public health response through dialogue and building public trust.

We have seen in previous disease outbreaks, and during the COVID-19 pandemic, how a fear-based or nonchalant narrative can be disastrous for an effective response.^46-49^ This highlights the critical role that communication plays in public health.^50^ There is more to learn about how the discourse around COVID-19 influences the pandemic response, however the impacts of this pandemic are far-reaching and far from over. We specifically illustrate important differences around gender, which should be further explored in future studies and related to effective response. As our world becomes more globalized and public health matters affect all of us, this era will reshape how public facing officials discuss both response and recovery policies. As a result, there is a need for more robust studies on communications in public health as we consider how the way we communicate will affect vaccine deployment, economic and social recovery, and more. While it has been overlooked in the past, this study contextualizes the importance of language, through a gender lens, in building both trust and an effective public health response.

## Author Contributions

SD developed the study methodology with the guidance of RD and collected the data for this study. SD and HA conducted the analysis and wrote the initial draft, with key input from MJB. RD provided edits and feedback. All authors revised the manuscript before submission

## Funding

This research received no specific grant from any funding agency in the public, commercial, or not-for-profit sectors.

## Competing Interests

We wish to confirm that there are no known conflicts of interest associated with this publication and there has been no financial support for this work that could have influenced its outcome. We confirm that this body of work has not been published elsewhere, nor is it currently under consideration for publication elsewhere.

## Data Availability

All data is available in the public domain but transcripts can be shared upon request.

## Acknowledgments

The authors would like to acknowledge Salla Atkins (Tampere University, Finland), Veronica Velasquez (Northwestern Trauma and Surgical Initiative, USA), Kaushi Ratnayake (Harper and Keele Veterinary School, UK), Alina Sobitschka (University of Göttingen, Germany), Andreia Bruno-Tomé (Monash University, Australia), Mehr Manzoor (Tulane University, USA), Daniela Suarez-Rebling (Icahn School of Medicine at Mount Sinai, USA) for transcription and/or translation assistance. We would also like to thank Rick Zednik (Women Political Leaders) and Emma Kinloch (King’s College London, UK) for their feedback on methodology and approach, Ann Keeling (Women in Global Health) for her review of the manuscript, and Lilly Khorsand (Women in Global Health) for assisting with table graphics.

---

**Appendix Table 1:** Inclusion/Exclusion of countries

| **COUNTRY** | **Leader** | **M/W** | **inclusion Rationale** | **Region** | **reason for exclusion** |
| --- | --- | --- | --- | --- | --- |
| ***Belgium*** | Wilmès | W | G10, UNSC, Woman | Europe | Included |
| ***France*** | Macron | M | G10, UNSC | Europe | Included |
| ***Germany*** | Merkel | W | G10, UNSC, Woman | Europe | Included |
| ***United Kingdom*** | Johnson | M | G10, UNSC | Europe | Included |
| ***United States*** | Trump | M | G10, UNSC | North America | Included |
| ***Brazil*** | Bolsonaro | M | BRICS | South America | Included |
| ***Russia*** | Putin | M | BRICS, UNSC | Europe/Asia | Included |
| ***India*** | Modi | M | BRICS | Asia | Included |
| ***South Africa*** | Ramaphosa | M | BRICS, UNSC | Africa | Included |
| ***Dominican Republic*** | Medina | M | UNSC | Caribbean | Included |
| ***Indonesia*** | Widodo | M | UNSC | Asia | Included |
| ***Niger*** | Issoufou | M | UNSC | Africa | Included |
| ***Saint Vincent and the Grenadines*** | Gonsalves | M | UNSC | Caribbean | Excluded – Speeches not available on public domain |
| ***Tunisia*** | Fakhfakh | M | UNSC | Africa | Excluded – Speech transcripts not available, no capacity to transcribe Arabic |
| ***Vietnam*** | Nguyễn Phú Trọng | M | UNSC | Asia | Excluded – Speech transcripts not available, no capacity to transcribe Vietnamese |
| ***Bangladesh*** | Hasina | W | Woman | Asia | Included |
| ***Barbados*** | Mottley | W | Woman | Caribbean | Excluded – Only 1 speech available, other Caribbean nation chosen |
| ***Bolivia*** | Añez | W | Woman | South America | Included |
| ***Denmark*** | Frederiksen | W | Woman | Europe | Excluded – Only 1 speech available, other European countries already represented |
| ***Finland*** | Marin | W | Woman | Europe | Included |
| ***Iceland*** | Jakobsdottir | W | Woman | Europe | Excluded – Speeches not available on public domain |
| ***Namibia*** | Kuugongelwa | W | Woman | Africa | Excluded – Speeches not available on public domain |
| ***New Zealand*** | Ardern | W | Woman | Oceania | Included |
| ***Norway*** | Solberg | W | Woman | Europe | Included |
| ***Scotland*** | Sturgeon | W | Woman | Europe | Included |
| ***Serbia*** | Brnabic | W | Woman | Europe | Speeches not available on public domain |
| ***Sint Maarten*** | Jacobs | W | Woman | Caribbean | Included |
| ***Taiwan*** | Ing-wen | W | Woman | Asia | Included |

**Appendix Table 2:** COVID-19 context

| Country | Leader | M/W | Total Population<sup>51</sup> | Confirmed Cases 29 Feb<sup>52</sup> | Cumulative Deaths<sup>52</sup> | Confirmed Cases 31 March<sup>53</sup> | Cumulative Deaths<sup>53</sup> | Confirmed Cases as 30 April<sup>54</sup> | Cumulative Deaths<sup>54</sup> | Confirmed Cases 31 May<sup>55</sup> | Cumulative Deaths<sup>55</sup> | Confirmed Cases 30 June<sup>25</sup> | Cumulative Deaths<sup>25</sup> | Cases Per Capita | Deaths Per Capita |
| --- | --- | --- | --- | --- | --- | --- | --- | --- | --- | --- | --- | --- | --- | --- | --- |
| **Brazil** | Bolsonaro | M | 212,559,417 | 1 | 0 | 4256 | 136 | 71886 | 5017 | 465166 | 27878 | 1344143 | 57622 | **0.6324%** | **0.0271%** |
| **Dominican Republic** | Medina | M | 10,847,910 | 0 | 0 | 901 | 320 | 6652 | 293 | 16908 | 498 | 31816 | 733 | **0.2933%** | **0.0068%** |
| **France** | Macron | M | 65,273,511 | 57 | 2 | 43977 | 3017 | 127066 | 24054 | 148436 | 28717 | 156930 | 29739 | **0.2404%** | **0.0456%** |
| **India** | Modi | M | 1,380,004,385 | 3 | 0 | 1071 | 29 | 33050 | 1074 | 182143 | 5164 | 566840 | 16893 | **0.0411%** | **0.0012%** |
| **Indonesia** | Widodo | M | 273,523,615 | 0 | 0 | 1414 | 122 | 9771 | 784 | 25773 | 1573 | 55092 | 1783 | **0.0201%** | **0.0007%** |
| **Niger** | Issoufou | M | 24,206,644 | 0 | 0 | 20 | 3 | 713 | 32 | 956 | 64 | 1075 | 67 | **0.0044%** | **0.0003%** |
| **Russia** | Putin | M | 145,934,462 | 2 | 0 | 1837 | 9 | 106498 | 1073 | 405843 | 4693 | 647849 | 9320 | **0.4439%** | **0.0064%** |
| **South Africa** | Ramaphosa | M | 59,308,690 | 0 | 0 | 1326 | 3 | 5350 | 103 | 30967 | 643 | 144264 | 2529 | **0.2432%** | **0.0043%** |
| **UK** | Johnson | M | 67,886,011 | 20 | 0 | 22145 | 1408 | 165225 | 26097 | 272840 | 38376 | 311969 | 43575 | **0.4595%** | **0.0642%** |
| **US** | Trump | M | 331,002,651 | 62 | 0 | 140640 | 2398 | 1003974 | 5428 | 1716978 | 101567 | 2537636 | 126203 | **0.7667%** | **0.0381%** |
| **Bangladesh** | Hasina | W | 164,689,383 | 0 | 0 | 49 | 5 | 7103 | 163 | 44608 | 610 | 141801 | 1783 | **0.0861%** | **0.0011%** |
| **Belgium** | Wilmès | W | 11,589,623 | 1 | 0 | 11899 | 513 | 47859 | 7501 | 58186 | 9453 | 61427 | 9747 | **0.5300%** | **0.0841%** |
| **Bolivia** | Añez | W | 11,673,021 | 0 | 0 | 97 | 5 | 1053 | 55 | 8731 | 300 | 31524 | 1014 | **0.2701%** | **0.0087%** |
| **Finland** | Marin | W | 5,540,720 | 2 | 0 | 1313 | 13 | 4906 | 206 | 6826 | 316 | 7209 | 328 | **0.1301%** | **0.0059%** |
| **Germany** | Merkel | W | 83,783,942 | 57 | 0 | 61913 | 583 | 159119 | 6288 | 181482 | 8500 | 194259 | 8973 | **0.2319%** | **0.0107%** |
| **New Zealand** | Ardern | W | 4,822,233 | 1 | 0 | 600 | 1 | 1129 | 19 | 1154 | 22 | 1178 | 22 | **0.0244%** | **0.0005%** |
| **Norway** | Solberg | W | 5,421,241 | 6 | 0 | 4226 | 26 | 7667 | 202 | 8411 | 236 | 8855 | 249 | **0.1633%** | **0.0046%** |
| **Scotland<sup>56</sup>** | Sturgeon | W | 5,254,800 | 0 | 0 | 1993 | 69 | 11353 | 1475 | 15400 | 2362 | 18251 | 2485 | **0.3473%** | **0.0473%** |
| **Sint Maarten** | Jacobs | W | 42,876 | 0 | 0 | 6 | 0 | 75 | 13 | 78 | 15 | 77 | 15 | **0.1796%** | **0.0350%** |
| **Taiwan** | Ing-wen | W | 23,816,775 | 39 | 1 | 322 | 5 | 429 | 6 | 442 | 7 | 447 | 7 | **0.0019%** | **0.0000%** |

## Notes

### Competing Interest Statement

The authors have declared no competing interest.

### Funding Statement

No external funding was received to conduct this research

